# Comparative effectiveness of allocation strategies of COVID-19 vaccines and antivirals against emerging SARS-CoV-2 variants of concern in East Asia and Pacific region

**DOI:** 10.1101/2021.10.20.21265245

**Authors:** Kathy Leung, Mark Jit, Gabriel M Leung, Joseph T Wu

## Abstract

**Background:** We aimed to evaluate the impact of various allocation strategies of COVID-19 vaccines and antiviral such that the pandemic exit strategy could be tailored to risks and preferences of jurisdictions in the East Asia and Pacific region (EAP) to improve its efficiency and effectiveness.

**Methods:** Vaccine efficacies were estimated from the titre distributions of 50% plaque reduction neutralization test (PRNT50), assuming that PRNT50 titres of primary vaccination decreased by 2-10 folds due to antibody waning and emergence of VOCs, and an additional dose of vaccine would increase PRNT50 titres by 3- or 9-fold. We then used an existing SARS-CoV-2 transmission model to assess the outcomes of vaccine allocation strategies with and without the use of antivirals for symptomatic patients in Japan, Hong Kong and Vietnam.

**Findings:** Increasing primary vaccination coverage was the most important contributing factor in reducing the total and peak number of COVID-19 hospitalizations, especially when population vaccine coverage or vaccine uptake among older adults was low. Providing antivirals to 50% of symptomatic infections only further reduced total and peak hospitalizations by 10-13%. The effectiveness of an additional dose of vaccine was highly dependent on the immune escape potential of VOCs and antibody waning, but less dependent on the boosting efficacy of the additional dose.

**Interpretation:** Increasing primary vaccination coverage should be prioritised in the design of allocation strategies of COVID-19 vaccines and antivirals in the EAP region. Heterologous vaccination with any available vaccine as the additional dose could be considered when planning pandemic exit strategies tailored to the circumstances of EAP jurisdictions.

**Funding:** Health and Medical Research Fund, General Research Fund, AIR@InnoHK

## Introduction

By implementing public health measures rapidly and effectively, most countries in the East Asia and Pacific region (EAP) kept COVID-19 cases and deaths much lower than in the US, Europe, and South America in 2020. However, the emergence of variants of concern (VOCs), especially the Delta variant which is more transmissible and has greater immune escape potential, changed the game the put stress on the EAP region in 2021. Many low- and middle-income countries in EAP were left behind to develop or purchase COVID-19 vaccines and antivirals. By mid-August 2021, more than half of the populations in the United States, United Kingdom, and more than a dozen of high-income and upper-middle-income countries have been fully vaccinated according to the initially designed immunisation schedule (mostly with two doses); while conversely, only 1.2% of population in low-income countries have received at least one dose ^1^. Molnupiravir, the antiviral that was reported to reduce the risk of COVID-19 hospitalization and death by about 50% in the phase 3 trial ^2^, costs $700 per five-day course which means it might not be available in most low- and middle-income countries in the short term.

Moreover, Delta has caused large outbreaks in highly vaccinated populations ^3^ and it has led to calls for an additional dose of COVID-19 vaccine, e.g. a third dose for most vaccines and a second dose for some adeno-vectored vaccines. On one hand, “breakthrough” infections and reduction of *in vitro* serum neutralizing antibodies over time have been observed ^4^, suggesting a potential waning in vaccine efficacy against infection; on the other hand, the severity of disease in vaccinated individuals is nevertheless much reduced compared to unvaccinated individuals, indicating that the vaccines are still highly effective in reducing hospitalizations and deaths ^5,6^.

While equity is important in health decision making, EAP jurisdictions would have various expectation of vaccines and antivirals due to differences in the society and economy. The first group of jurisdictions, which has pursued a Zero-COVID or elimination strategy to stringently control COVID-19 within their borders, includes mainland China, Hong Kong, Macau, and Taiwan. A vaccine-driven exit strategy is the most likely path for these jurisdictions: if they were to minimise infections of VOCs in a highly susceptible population in the absence of public health measures, higher protection from vaccines would be required, which might only be attained by fully vaccinating all the eligible individuals with both primary and additional doses of the most efficacious vaccines ^7^. The second group of jurisdictions, which is in transition to a Living-with-COVID strategy, includes Australia, Japan, Singapore, South Korea, and most of the Southeast Asian countries. If their goals were to minimise COVID-19 hospitalizations to avoid overwhelming the health system, additional doses of vaccines might not be as necessary if the primary vaccination coverages are high and sufficient supplies of antivirals are secured. Furthermore, vaccine effectiveness also depends on the type of vaccines available, age distribution and underlying health condition of vaccinees and the population, emergence of new VOCs and waning of protection over time ^8^.

Therefore, it is important to evaluate the impact of various allocation strategies of vaccines and antiviral such that the pandemic exit strategy could be tailored to jurisdictions’ risks and preferences to improve its efficiency and effectiveness. Here, we used an existing SARS-CoV-2 transmission model to assess the outcomes of primary and additional vaccine allocation strategies with and without the presence of antivirals. We defined primary vaccination as completion of the initially designed immunisation schedule, which consisted of two doses for most vaccines and one dose for some adeno-vectored vaccines. Vaccines that were given beyond primary vaccination were defined as additional vaccination. We assumed primary vaccination, i.e., vaccinating the never-vaccinated as much as possible, should be always prioritized. Given that the vaccines that many EAP countries receive from the COVAX initiative compose of different types, we analysed hypothetical jurisdictions which mainly used 1) inactivated virus vaccines, 2) mRNA vaccines, 3) adeno-vectored vaccines and 4) a mix of inactivated/adeno-vectored virus and mRNA vaccines, with and without antivirals.

## Methods

### Predicting the reduction in vaccine efficacy due to emerging VOCs

Currently, three types of vaccines are most widely used in the world, including the adenovirus vector vaccines (e.g. AZD1222 from AstraZeneca), mRNA vaccines (e.g. BNT162b2 from BioNTech) and inactivated virus vaccines (e.g. CoronaVac from Sinovac) ^8^. Data from a cohort study of 49 BNT162b2 vaccinees and 49 CoronaVac vaccinees in Hong Kong showed that both vaccines elicited significant immune response against the original virus ^9^. At one month after the second dose of vaccine, the logarithmic of 50% plaque reduction neutralization test (PRNT50) titres among vaccinees follow a normal distribution, with the mean titre against the original virus of 251.60 (±1SD: 146.66 – 431.63) among BNT162b2 vaccinees and 69.45 (±1SD: 28.07 – 171.77) among CoronaVac vaccinees respectively (Figure S1).

Although comparable data about the AZD1222 vaccinees were not available in Hong Kong, it has been reported that the mean titres of pseudovirus and live virus neutralization tests among AZD1222 vaccinees were 135-172 and 166-261 respectively ^10^, and the efficacies of AZD1222 vaccines against the original virus lied between CoronaVac and BNT162b2 vaccines ^11^. Therefore, without loss of generalization, we assumed that the mean PRNT50 titre against the original virus of AZD1222 vaccinees were 133.82 (±1SD: 78.59 –227.85), which was the average of CoronaVac and BNT162b2 vaccinees in the log scale.

We assumed that vaccine efficacies were reduced mainly due to antibody waning and emergence of VOCs ^6,12-14^. Comprehensive analyses of neutralization using 16 isolates of distinct SARS-CoV-2 variants revealed a range of reduction in the neutralization titres among vaccinees who were previously not infected, with an average of 0.5-3.4, 1.6-6.9 and 2.3-13.2 folds of decrease in neutralization titre for the Alpha (B.1.1.7), Delta (B.1.617.2) and Beta (B.1.351) variants, varied by the time since the second dose of mRNA vaccines ^4,15^. As such, we estimated the vaccine efficacy against infections (i.e., *σ*_*m*_) of emerging VOCs by reducing the mean PRNT50 titre level against the original virus strain by a factor of 2, 4, 7, or 10 for hypothetical variants of concern (see Appendix for details).

### Assumption about efficacy of vaccines and antivirals

The analyses above estimated vaccine efficacy against SARS-CoV-2 infection. However, it is thought that the immune response after vaccination, especially cellular immunity, may provide greater protection from severe disease than from mild or asymptomatic infection ^16^. Khoury et al estimated that the neutralization titre level for 50% protection from infection was comparable to the level required for about 85% protection from severe disease ^17^. Moreover, T cell responses or reactivation of memory B cell responses may also play an important role in protection from severe diseases ^16,18-20^. Therefore, we assumed that the vaccine efficacy in reducing symptomatic diseases and admission to hospital (i.e., *σ*_*S*_) was 25% higher than *σ*_*m*_, i.e., *σ*_*S*_ = 1.25 × *σ*_*m*_ and fell within the range between 65% and 95% for emerging VOCs, given that there were limited data about the association between T cell response level and *σ*_*s*_. Our assumption about *σ*_*S*_ was more conservative compared with the observed data on the existing VOCs for most vaccines ^13,16^. Similarly, we assumed that the vaccine efficacy in reducing infectiousness if infected (i.e., *σ*_*t*_) was 20% lower than *σ*_*m*_, i.e., *σ*_*t*_ = 0.8 × *σ*_*m*_, for variants of concern, following similar assumptions used by Scientific Advisory Group for Emergencies of the UK ^21^. For antivirals, we assumed the efficacies were *ε*_*m*_ = 0 in reducing susceptibility to infection, *ε*_*t*_ = 0 in reducing infectivity if infected and *ε*_*S*_ = 0.5 in reducing symptomatic diseases and hospitalizations, since molnupiravir is currently only recommended to patients within 5 days of symptom onset ^2^.

### Modelling allocation strategies of COVID-19 vaccines and antivirals

We adapted an existing age-structured susceptible-infectious-removed model of SARS-CoV-2 transmission dynamics that can be parameterized with country- or region-specific age demographics and contact patterns to simulate the effect of vaccination (i.e., including primary vaccination and booster vaccination) on transmission ^7,22^.

In three hypothetical populations, we parameterized the model with population demographics and age-specific primary vaccine uptake as of 1 August 2021 from Japan (medium population coverage of 39.7% and high coverage of 87.6% among older adults aged 65 or above), Hong Kong (medium population coverage of 44.5% and low coverage of 19% among older adults aged 70 or above) and Vietnam (low population coverage of 6.0% and minimal coverage among older adults aged 65 or above), to assess the effectiveness of allocation strategies (Figure S2).

Then we assumed: 1) 100% of vaccinees received BNT162b2 vaccines in Japan, 2) 60% and 40% of vaccinees received BNT162b2 and CoronaVac vaccines in Hong Kong, and 3) 40%, 30% and 30% of vaccinees received BNT162b2, AZD1222, CoronaVac vaccines in Vietnam. Since giving additional vaccines to vaccinees who have completed primary vaccination would have lower marginal effectiveness compared with vaccinating individuals who were never vaccinated (inferred from the titre distribution after the third dose ^23^), therefore, throughout the analysis, we hypothesised that primary vaccination should always be prioritised and considered the following vaccine allocation strategies:

#### Strategy 1: Vaccinate the never-vaccinated individuals only as much as possible (PV)

Considering the potential vaccine hesitancy and refusal, we assumed that the maximum primary vaccination coverage would achieve 70%, 80% or 90% in three scenarios.

#### Strategy 2: Vaccinate the never-vaccinated individuals only as much as possible (PV) and provide antivirals to 50% of the symptomatic patients (PV+AR)

We assumed that the maximum primary vaccination coverage would achieve 70%, 80% or 90% in three scenarios. We assumed 40% of vaccinees (i.e., completed the primary vaccination) and 60% of unvaccinated individuals would develop symptoms if they were infected (Table S4), and antivirals would be provided to 50% of the symptomatic patients.

#### Strategy 3: Vaccinate the never-vaccinated individuals as much as possible, give an additional dose to only AZD1222 or CoronaVac vaccinees and provide antivirals to 50% of the symptomatic patients (PV+AV+AR)

We assumed that the maximum vaccination coverage would achieve 70%, 80% or 90% and that all AZD1222/CoronaVac vaccinees were willing to take the additional dose regardless of the type of vaccine that would be given. We assumed that the PRNT50 titres of AZD1222/CoronaVac vaccinees would increase by 3 or 9 folds after the additional dose, which was consistent with the findings in the recent studies about the immunogenicity of a third dose of AZD1222/CoronaVac vaccine (i.e. 2-4 folds increase in antibody titres and T cell response among AZD1222 vaccinees ^24^, and 3-4 folds increase of antibody titres in healthy adults aged 18 to 59 and ≥7 folds among the older adults aged 60 or above among CoronaVac vaccinees ^25,26^).

#### Strategy 4: Vaccinate the never-vaccinated individuals as much as possible, give an additional dose to all vaccinated individuals with PRNT50 titre lower than a certain threshold, and provide antivirals to 50% of the symptomatic patients (PV+ selective AV+AR)

Similarly, we assumed that the maximum vaccination coverage would achieve 70%, 80% or 90% and that all vaccinees were willing to take the additional dose if given. We assumed that the PRNT50 titre of BNT162b2 vaccinees would at least increase by 3 or 9 folds after the additional dose ^23^. The additional dose would only be given to vaccinees with PRNT50 titres lower than the prespecified thresholds. We considered three scenarios in which the additional dose would be given to vaccinees with the original PRNT50 titre lower than 25.9, 39.9 and 74.1, which corresponded to the estimated thresholds of 50%, 60% and 70% protection against infection of the original virus..

#### Strategy 5: Vaccinate the never-vaccinated individuals as much as possible, give an additional dose to all vaccinated individuals and provide antivirals to 50% of the symptomatic patients (PV+ universal AV+AR)

We assumed that the maximum vaccination coverage would achieve 70%, 80% or 90% and that all vaccinees were willing to receive the additional dose. We assumed both additional dose of homologous and heterologous vaccines would elicit similar increases in PRNT50 titres, though only limited data were currently available from selected patient groups ^27^.

### Comparative effectiveness of allocation strategies

Similar to our previous analysis ^7^, we assumed that vaccines were first allocated to never vaccinated individuals in proportion to the age-specific vaccine uptake as of 1 August 2021 until the population vaccination coverage achieved 70%, 80% or 90% or the vaccine uptake of a specific age group achieved 100%. Then the additional doses were allocated to vaccinees following the five allocation strategies above.

We assumed that the population was completely susceptible before vaccination, i.e., the population immunity was only obtained from vaccination. In view of the emergence of VOCs, we assumed the pre-vaccination effective reproductive (*R*_*e*_) was 6 even with limited public health and social measures in place (e.g. the basic reproductive number of the Delta variant was assumed to be 6, i.e., ∼300% higher than the original virus strain ^28^). We then simulated the epidemics and estimated the total number of hospitalizations and the maximum daily number of hospitalizations.

We estimated the total number of vaccine doses required for every COVID-19 hospitalization averted in Strategy 1-5 compared with the existing vaccination uptake as of 1 August 2021 (Strategy 0). We also estimated the number of primary and additional vaccine doses required for every COVID-19 hospital admission averted in Strategy 1-5, compared with Strategy 0. Other parameter values were shown in the Appendix.

## Results

### Reduction in vaccine efficacy due to antibody waning and VOCs

We estimated that vaccine efficacy in reducing susceptibility (*σ*_*m*_) to infection decreased substantially due to VOCs and antibody waning (Figure 1): *σ*_*m*_ of BNT162b2 vaccinees decreased from 89% to 84%, 68%, 45% and 28% when PRNT50 titres decreased by 2, 4, 7 and 10 folds; *σ*_*m*_ of AZD1222 vaccinees decreased from 85% to 70%, 41%, 15% and 6%; and *σ*_*m*_ of CoronaVac vaccinees decreased from 65% to 42%, 20%, 8% and 4% respectively. Similarly, we estimated *σ*_*t*_ (i.e., vaccine efficacy in reducing infectiousness) decreased from 72% to 67 %, 54%, 36%, 23% for BNT162b2 vaccinees, from 68% to 56%, 33%, 12% and 5% for AZD1222 vaccinees, and from 52% to 33%, 16%, 7% and 3% for CoronaVac vaccinees. For vaccine efficacy in reducing symptomatic disease and hospitalizations, we estimated *σ*_*S*_ reduced from 95% to 95%, 85%, 65%, 65% for BNT162b2 vaccinees, from 95% to 87%, 65%, 65%, 65% for ADZ1222 vaccinees, and from 81% to 65%, 65%, 65%, 65% for CoronaVac vaccinees. The estimated vaccine efficacies were largely consistent with findings from case-control trials or real world observations about Alpha and Delta variants ^8^, for which PRNT50 titres were estimated to decreased by 1-4 folds and 2-7 folds respectively ^4^.

**Figure 1.**
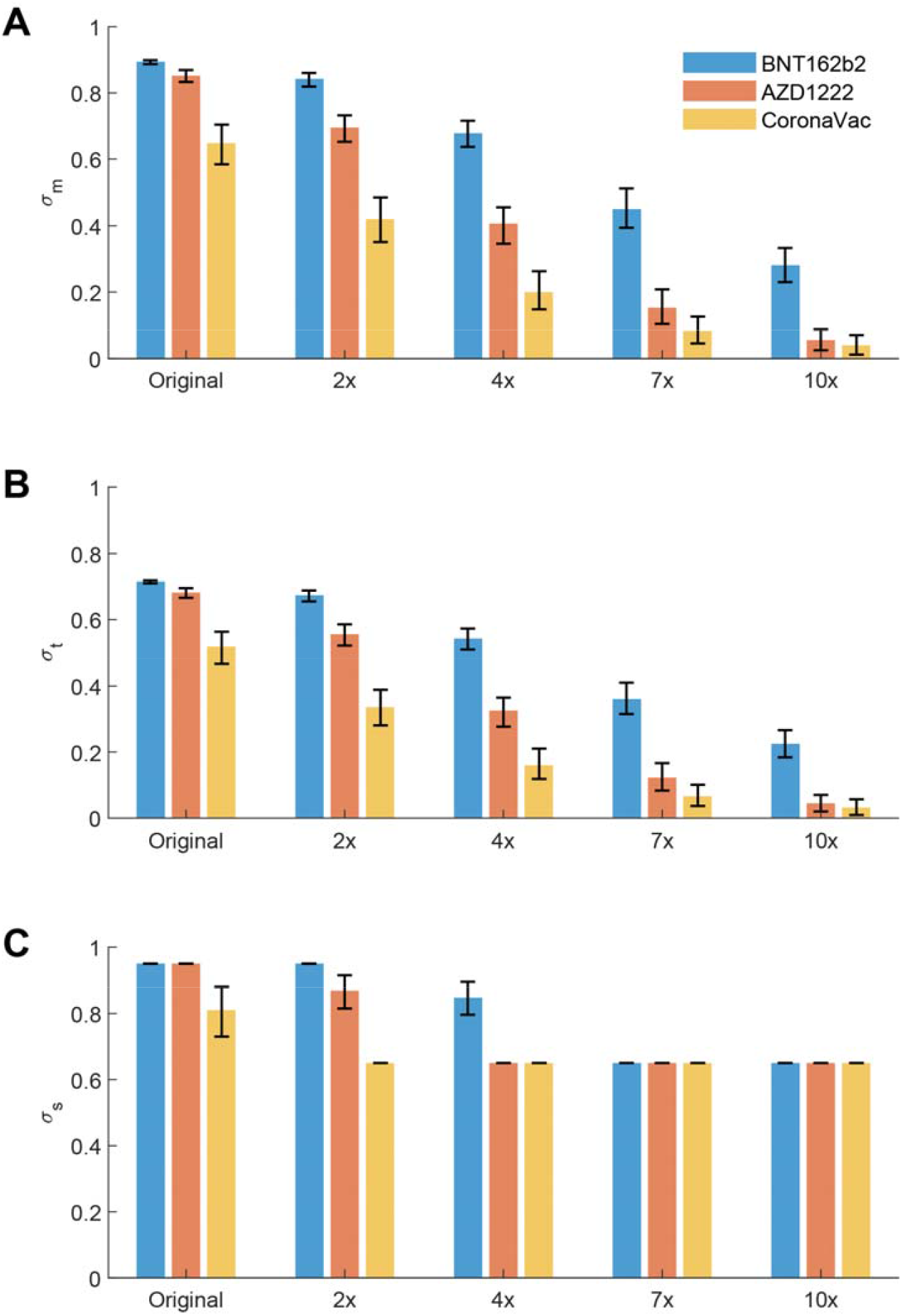
Estimated vaccine efficacies of BNT162b2, AZD1222 and CoronaVac vaccines from PRNT50 titre distributions. The mean PRNT50 titres of vaccinees against the hypothetical variants of concern were assumed to decrease by 2, 4, 7, and 10 folds respectively compared with the PRNT50 titres against the original virus strain. The vaccine efficacies were estimated by bootstrapping the PRNT50 titres of 100 vaccinees by 1000 times. The error bars showed the 95% CI of the estimates with the uncertainty from PRNT50 titre distributions. (A) Vaccine efficacy in reducing susceptibility to infection (*σ*_*m*_). (B) Vaccine efficacy in reducing infectiousness if infected (*σ*_*t*_). (C) Vaccine efficacy in reducing symptomatic disease and hospital admission (*σ*_*S*_).

### Effectiveness of allocation strategies of vaccines and antivirals

We assessed the effectiveness of allocation strategies of vaccines and antivirals in Japan, Hong Kong, and Vietnam (Figure 2-4). As expected, increasing primary vaccination coverage was the most important contributing factor in reducing the total and peak number of COVID-19 hospitalizations in Strategy 1-5 (Figure 2-4), especially when the population vaccine coverage or the vaccine uptake among older adults was low (Figure 2-3). In Vietnam, increasing primary vaccination coverage from 6% to 70%, 80%, 90% reduced the total hospitalizations by 53-62%, 64-76%, 64-77% in Strategy 1 with no additional dose or antivirals when PRNT50 titres against VOCs reduced by 4-10 folds.

**Figure 2.**
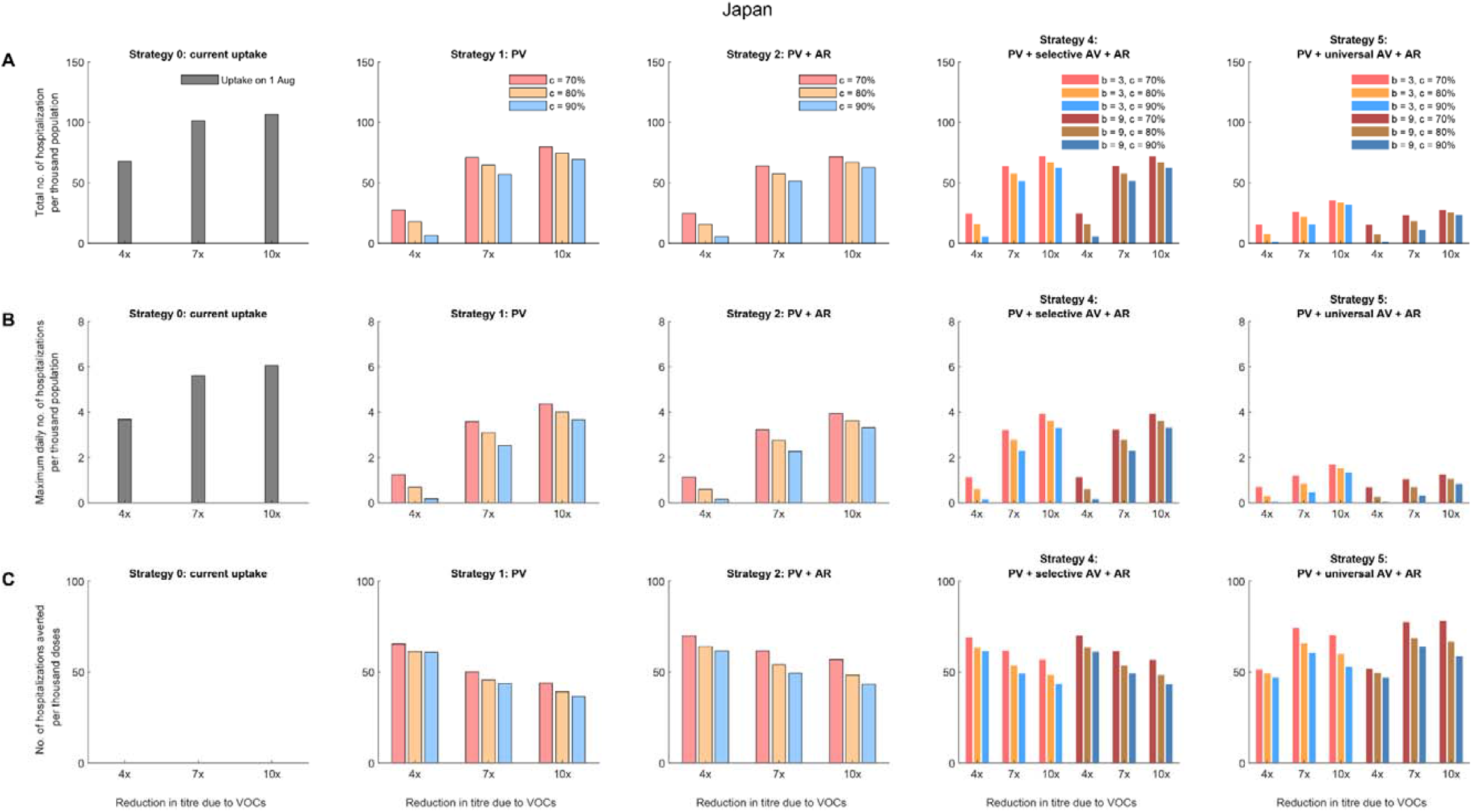
Comparative effectiveness of allocation strategies of vaccines and antivirals in Japan assuming all vaccinees received the BNT162b2 vaccines. As of 1 August 2021, 39.7% of the population had taken at least first dose of vaccine and all of them received the BNT162b2 vaccines. We assumed that the effective reproductive number before the vaccination program (*R*_*e*_) was 6 and that the effects of vaccination (characterized by *σ*_*m*_, *σ*_*t*_, and *σ*_*s*_) were realized instantaneously after the target vaccine uptake was achieved. For Strategy 4, the additional dose would be given to vaccinees with the original PRNT50 titre lower than 74.1, which corresponded to the estimated thresholds of 70% protection against the original virus. The epidemics were seeded with one introduction event immediately after the target vaccine uptake was achieved. We assumed PRNT50 titres reduced by 4, 7 or 10 folds due to VOCs and one dose of booster vaccine increased the PRNT50 titres by 3 or 9 folds (i.e., *b* = 3 or 9). We assumed the target vaccination coverage (*c*) was 70%, 80% or 90%. (A) The total number of hospitalizations per thousand population. (B) The maximum daily number of hospitalizations per thousand population. (C) The number of hospitalizations averted per thousand vaccine doses, compared with the existing vaccine uptake as of 1 August 2021 (i.e., about 39.7% coverage).

**Figure 3.**
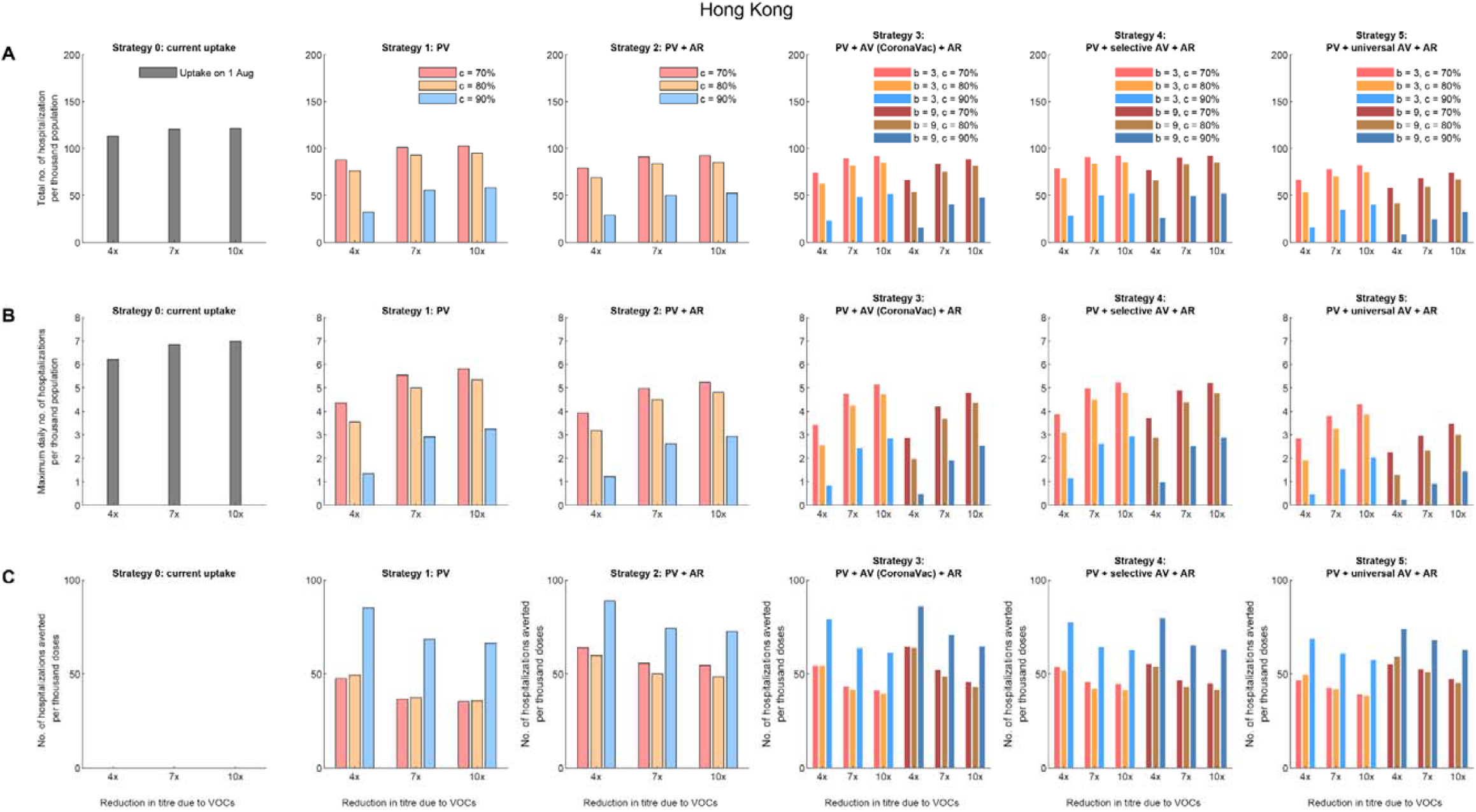
Comparative effectiveness of allocation strategies of vaccines and antivirals in Hong Kong assuming 60% vaccinees received the BNT162b2 vaccines and 40% received CoronaVac vaccines. (A) The total number of hospitalizations per thousand population. (B) The maximum daily number of hospitalizations per thousand population. (C) The number of hospitalizations averted per thousand vaccine doses, compared with the existing vaccine uptake as of 1 August 2021 (i.e., about 44.5% coverage).

**Figure 4.**
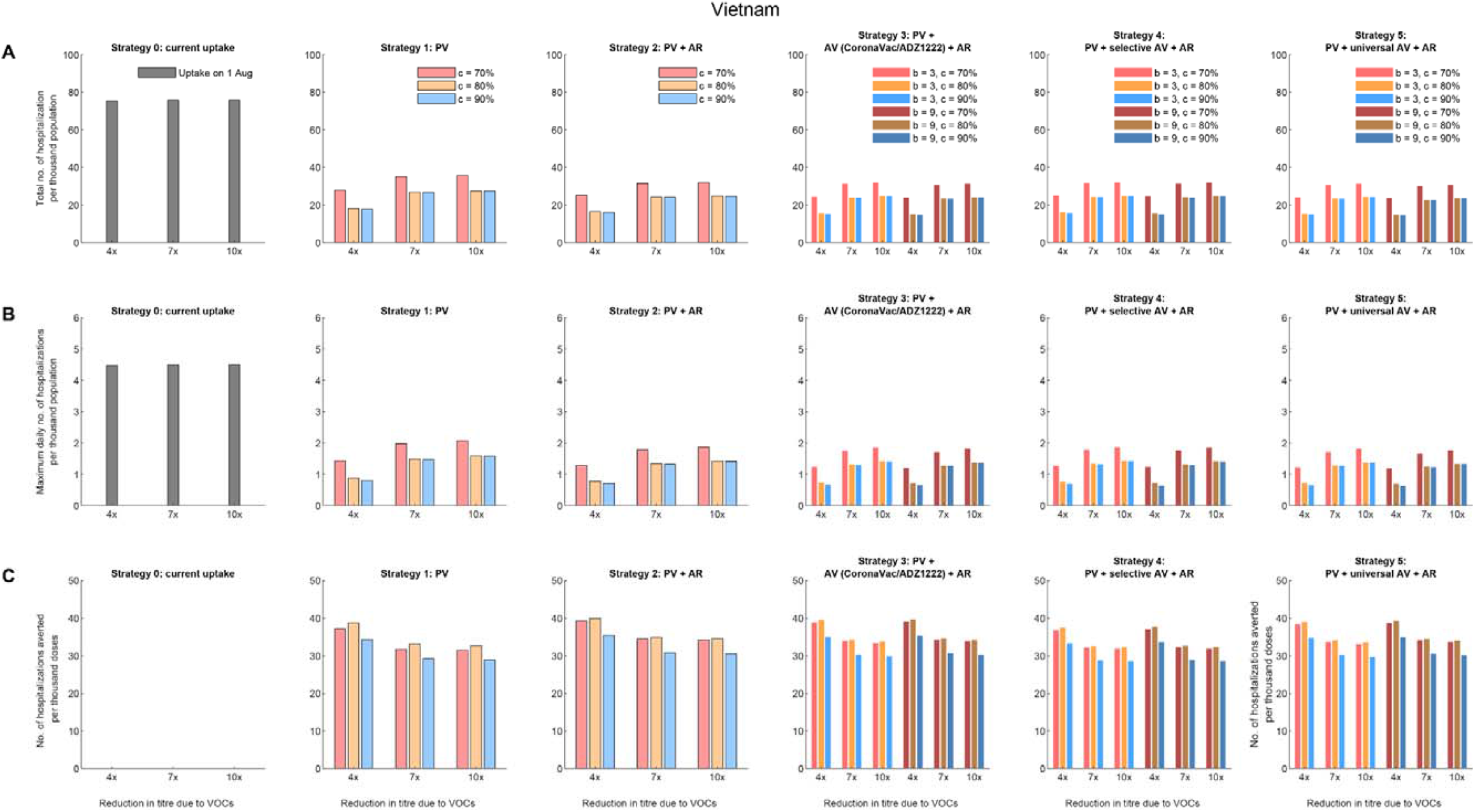
Comparative effectiveness of allocation strategies of vaccines and antivirals in Vietnam assuming 40%, 30% and 30% of vaccinees received BNT162b2, ADZ1222 and CoronaVac vaccines respectively. (A) The total number of hospitalizations per thousand population. (B) The maximum daily number of hospitalizations per thousand population. (C) The number of hospitalizations averted per thousand vaccine doses, compared with the existing vaccine uptake as of 1 August 2021 (i.e., about 6% coverage).

Similar proportions of 54-68%, 65-80%, 65-82% were reduced in peak hospitalisation accordingly. Similarly, in Hong Kong, increasing primary vaccination coverage from 44.5% to 70%, 80%, 90% reduced the total hospitalizations by 15-22%, 22-32%, 52-72% in Strategy 1 with no additional dose or antivirals and the peak hospitalizations reduced by 17-30%, 24-43%, 54-78%.

Compared with Strategy 1, providing antivirals to 50% of symptomatic infections in Strategy 2 only further reduced total and peak hospitalizations by 10-13% and 10-12% in Japan, Hong Kong, and Vietnam across all scenarios.

Across Strategy 3-5, the effectiveness of an additional dose of vaccine was highly dependent on the immune escape potential of VOCs or antibody waning. The total and peak hospitalizations increased with the decrease in PRNT50 titres due to VOCs. However, an additional dose of vaccine that generated higher increase in PRNT50 titres would not substantially increase the impacts of the vaccination program in Strategy 3 and 4 unless the additional dose was given universally to all vaccinees in Strategy 5: the total and peak hospitalizations, and the number of hospitalizations averted per thousand doses were not significantly different for additional dose that increased PRNT50 titres by 4, 7, and 10 folds in Strategy 3 and 4, whilst Strategy 5 had the largest impact in reducing total and peak hospitalizations in Japan with a high starting primary vaccine uptake among the elderly (Figure 2).

### Impacts of different age-specific vaccine uptakes

Across all strategies, the number of hospitalizations averted per thousand doses of vaccines reduced with the increase of primary vaccine uptake from 70% to 90% in Japan (Figure 2 and Figure S3) and Vietnam (Figure 3 and Figure S6-S8). However, in contrast to Japan and Vietnam, the number of hospitalizations averted per thousand doses of vaccines increased significantly with the increase of primary vaccine uptake to 90% in Hong Kong (Figure 3 and Figure S4-S5), which started with a low uptake among the elderly population on 1 August 2021.

### Impacts of the increase in PRNT50 titres after the additional dose of vaccine

The impact of an additional dose of vaccine in Strategy 3-5 was small compared with the effects of increasing primary vaccine uptake, especially when the additional dose did not increase PRNT50 titres substantially. Even when the additional dose was given universally to all vaccinees, the impact was dependent on the increase in PRNT50 titres after receiving the additional dose only when a less efficacious vaccine was widely used in the primary vaccination (Figure S3 and Figure S5).

### Strategies tailored to the preference of EAP jurisdictions

Strategy 5, which gave the additional dose universally to all vaccinees and provided antivirals to 50% of symptoms, was the most effective in reducing the total and peak hospitalizations, especially when immune escape of VOCs did not result in substantial waning in PRNT50 titres. Facing the threat by Delta variant, Strategy 5 might be the only option for EAP jurisdictions aiming to retain Zero-COVID status. For example, in Hong Kong, the maximum daily number of hospitalizations per thousand population was 0.23 if the primary vaccine coverage reached 90%, the PRNT50 titres decreased by 4 folds due to VOCs, antivirals were given to 50% symptomatic patients, and an additional dose that increased PRNT50 titres by 9 folds was given universally to all vaccinees (Figure 3); and this translated to the peak daily number of hospitalizations of 1,725, which was 4.6 times more than the maximum daily number of hospitalizations that Hong Kong would be able to manage (Table S4). As such, even with Strategy 5, some flexible and short-term public health and social measures should be retained during the planning of pandemic exit strategy.

However, for EAP jurisdictions aiming for Living-with-COVID, especially those with high primary vaccination coverage across all age groups with an efficacious vaccine, Strategy 2-4 might be sufficient with no or few additional doses of vaccines. For example, in Japan, the maximum daily number of hospitalizations per thousand population was 0.15 if the primary vaccine coverage reached 90%, the PRNT50 titres decreased by 4 folds due to VOCs, and antivirals were given to 50% of symptomatic patients (Figure 2); and this translated to less than 3 times of the assumed health system capacity (Table S4). Given that Japan had experienced multiple waves of outbreaks and their health system is relatively resilient, additional doses of vaccines could be provided to only selected groups of the population.

## Discussion

Our modelling analyses provided evidence to support that increasing primary vaccination coverage was the most important contributing factor to reduce COVID-19 hospitalizations and deaths in the mass vaccination programmes. The effects of increasing primary vaccination coverage were most prominent when the vaccine uptake among older adults was low, such as in the population of Hong Kong, suggesting allocation strategies should prioritise protecting the most vulnerable groups to reduce COVID-19 disease burden through vaccination. Antivirals added minimal impacts in reducing total and peak hospitalizations, although we optimistically assumed 50% of symptomatic patients were provided with antivirals.

Our findings showed that selectively giving additional doses to individuals with lower antibody titres, such as AZD1222/CoronaVac vaccinees only in Strategy 3 or vaccinees with PRNT50 titre lower than a prespecified threshold in Strategy 4, added limited net benefits in terms of reducing the total and peak COVID-19 hospitalizations. Furthermore, when vaccine uptake was low among older adults or a less effective vaccine was widely used, additional doses of vaccines were more effective only when the primary vaccination coverage was higher (such as in Hong Kong in Figure 3 and Figure S5). Our analyses further supported that increasing primary vaccination coverage was essential and should be prioritised.

Interestingly, we found that the effectiveness of the additional dose was negatively associated with the potential immune escapes of VOCs but was less dependent on the boosting efficacy, i.e., the increase in antibody titres generated after the additional dose. Since nearly all COVID-19 vaccines available increased neutralizing antibody titres by at least four folds ^23,25,26^, heterologous vaccination with any available vaccines, as long as they are safe, could be potential options when planning the allocation strategy.

Our study has several limitations. First, we did not consider vaccine availability and the level of vaccine hesitancy in different populations. In Hong Kong, the low vaccine uptake among older adults is not because of limited vaccine supply but largely due to worries about vaccine safety among individuals of old age with comorbidities. Whereas for most Southeast Asian countries in the EAP region, increasing primary vaccine uptake to 90% might not be feasible in the short term. Second, our model assumed that the population was completely susceptible to SARS-CoV-2 and that there was no virus circulating before the commencement of vaccination programmes as a case study. Populations that have experienced multiple waves of COVID-19 previously might not need additional doses or antivirals. Recent studies found that natural infections with one dose of vaccines generated higher neutralizing titres and was correlated with highest efficacy against symptomatic infections and hospitalizations ^29^. Finally, we assumed that limited public health and social measures would be maintained after the primary and additional vaccinations, and a high starting effective reproductive number (*R*_*e*_ = 6) of VOCs in the absence of vaccination. Gradual relaxation of public health and social measures would leave more options for the allocation strategies of vaccines and antivirals.

In summary, our analyses provided support to the prioritization of increasing primary vaccination coverage. Heterologous vaccination with any available vaccines as the additional dose, as long as it is safe, could be considered as potential options when planning the pandemic exit strategies tailored to the preference of EAP jurisdictions.

## Data Availability

All data produced in the present work are contained in the manuscript.

## Data sharing statement

We collated all data from publicly available data sources. All data included in the analyses are available in the main text or the supplementary materials.

## Acknowledgement

We thank Shirley Kwok from School of Public Health, The University of Hong Kong for technical support.

## Contributors

KL, MJ, JTW and GML designed the study. KL, MJ, JTW and GML developed the model and analysed data. KL, MJ, JTW and GML interpreted the results and wrote the manuscript.

## Appendix

### Distribution of neutralizing antibody titres against the original virus among vaccinees in Hong Kong

Two vaccines are currently available in the COVID-19 vaccination program in Hong Kong, including an mRNA vaccine (BNT162b2; BioNTech-Fosun) an inactivated virus vaccine (CoronaVac; Sinovac). As of 1 August 2021, 50% of the population have received at least one dose of vaccine (Table S1), including 60% vaccinees who received the BNT162b2 vaccines and 40% vaccinees who received the CoronaVac vaccines ^30^. We next obtained data from a cohort study of 49 BNT162b2 vaccinees and 49 CoronaVac vaccinees ^9^. At one month after the second dose of vaccine, the logarithmic of 50% plague reduction neutralization test (PRNT50) titres among vaccinees follow normal distribution, with the mean titre against the original virus of 251.60 (±1SD: 146.66 – 431.63) among BNT162b2 vaccinees and 69.45 (±1SD: 28.07 – 171.77) among CoronaVac vaccinees respectively (Figure S1).

Although comparable data about the AZD1222 vaccinees were not available, it has been reported that the vaccine efficacies of AZD1222 against the original virus lies between CoronaVac and BNT162b2 vaccines ^11^. Therefore, without loss of generalization, we assumed the mean PRNT50 titre against the original virus of AZD1222 vaccinees were 133.82 (±1SD: 78.59 –227.85), which was the average of CoronaVac and BNT162b2 vaccinees in the log scale.

### Estimation of vaccine efficacy against infection from the neutralizing titre distribution

We adapted the predictive model developed by Khoury et al to estimate the immune protection levels against SARS-CoV-2 infection among vaccinees ^17^. With PRNT50 titres from 293 RT-PCR confirmed symptomatic and asymptomatic infections in Hong Kong ^31^, we estimated that the PRNT50 titre threshold for 50%, 60%, 70%, 80% and 90% protection against SARS-CoV-2 infection were approximately 25.9, 39.9, 54.4, 74.1 and 99.7. By comparing these protection titre thresholds with the PRNT50 titre distribution of vaccinees, we also estimated the proportion of BNT162b2 and CoronaVac vaccinees with PRNT50 titre at or above the thresholds (Table S2).

To model the relationship between neutralization titre distribution of vaccinees and the vaccine efficacy, let *F*(*r*) denote the cumulative density function of the PRNT50 titre distribution after vaccination where *r* denotes the PRNT50 titre threshold and *F*(*r*) denotes the proportion of vaccinees with PRNT50 titre at or above the threshold *r*. The probability density function of the PRNT50 titre distribution *f*(*r*) was estimated from *F*(*r*). Let *σ*_*m*_ (*r*) denote the efficacy in reducing susceptibility to infection among vaccinees with PRNT50 titre at or above the threshold *r*, and *ε*_*m*_ (*r*) be the vaccine efficacy among vaccinees with PRNT50 titre at the threshold *r*, such that 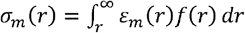 (Table S2 and S3). Similarly, we estimated *σ*_*m*_ among all vaccinees as 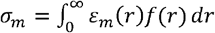.

**Figure S1.**
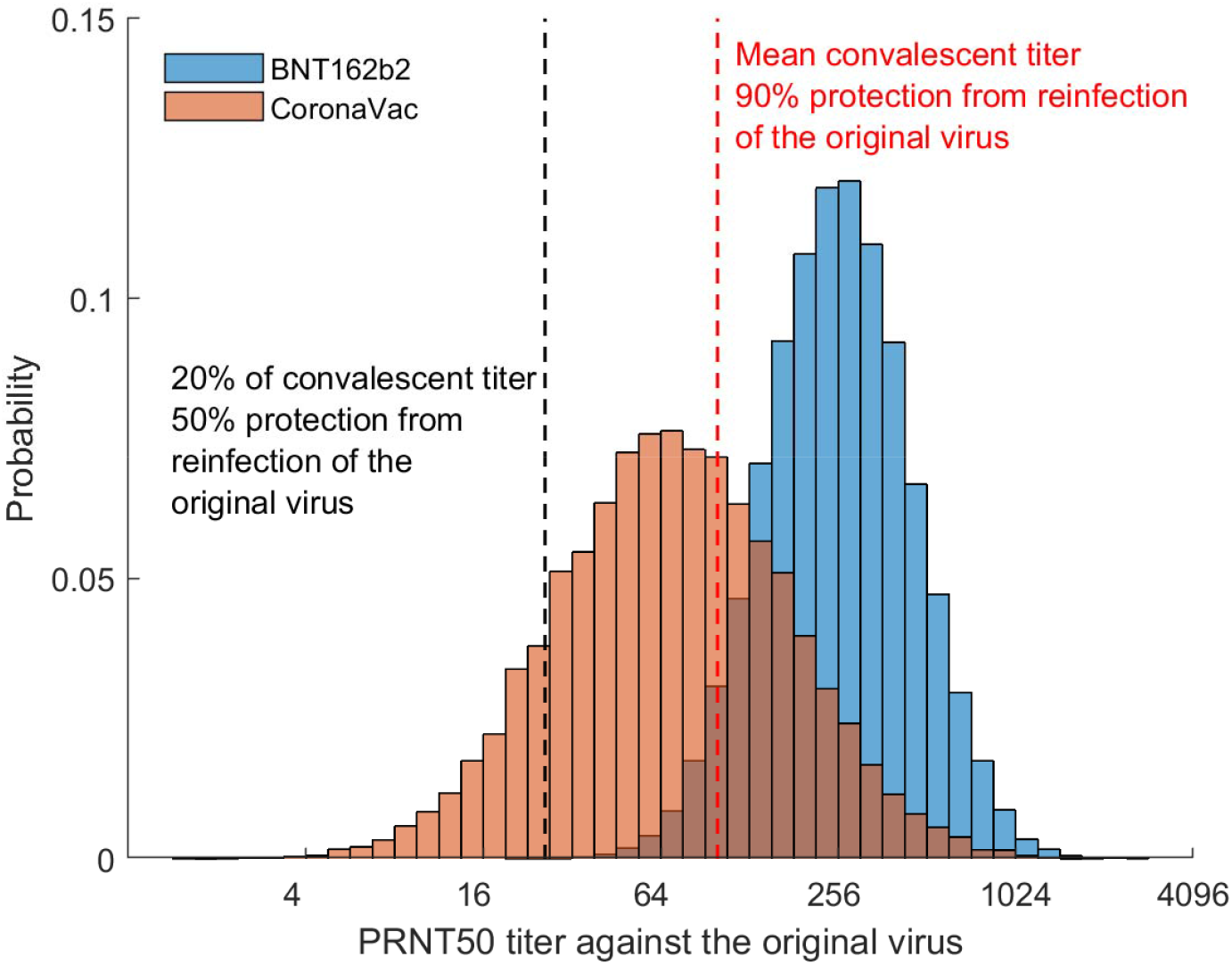
Distribution of PRNT50 titres against the original virus among vaccinees in Hong Kong. The distributions of BNT162b2 and CoronaVac vaccinees were estimated from the data presented in Mok et al ^9^. The black dashed line showed the estimated mean convalescent PRNT50 titre of infections based on Lau et al ^31^, which corresponds to 50% protection against infection of the original virus according to Khoury et al ^17^. Similarly, the red dashed line showed the estimated PRNT50 titre threshold for 90% protection against infection of the original virus.

**Figure S2.**
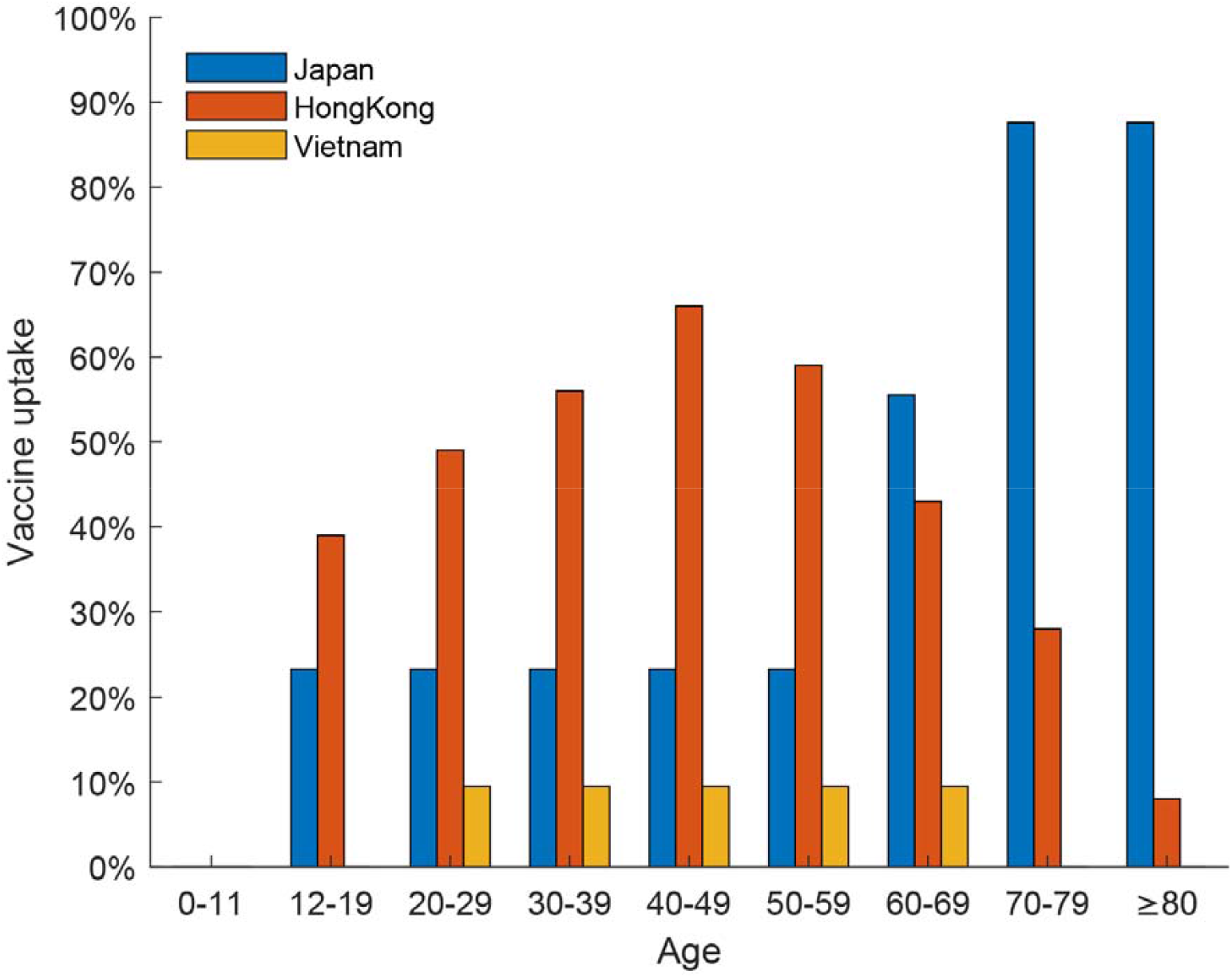
Age-specific vaccine uptake in Japan, Hong Kong, and Vietnam as of 1 August 2021.

**Figure S3.**
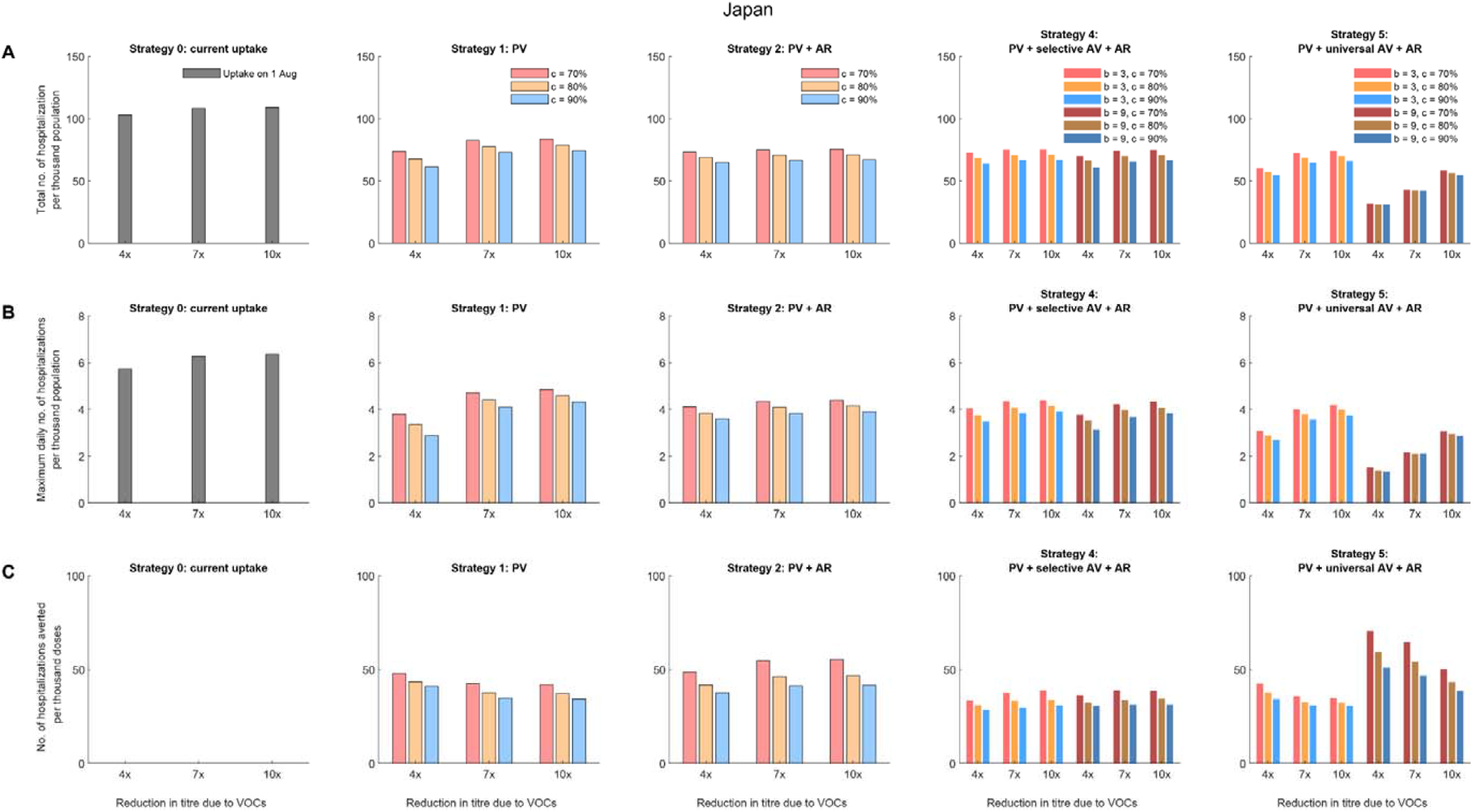
Comparative effectiveness of allocation strategies of vaccines and antivirals in Japan assuming all vaccinees received the CoronaVac vaccines. (A) The total number of hospitalizations per thousand population. (B) The maximum daily number of hospitalizations per thousand population. (C) The number of hospitalizations averted per thousand vaccine doses, compared with the existing vaccine uptake as of 1 August 2021 (i.e., 39.7%).

**Figure S4.**
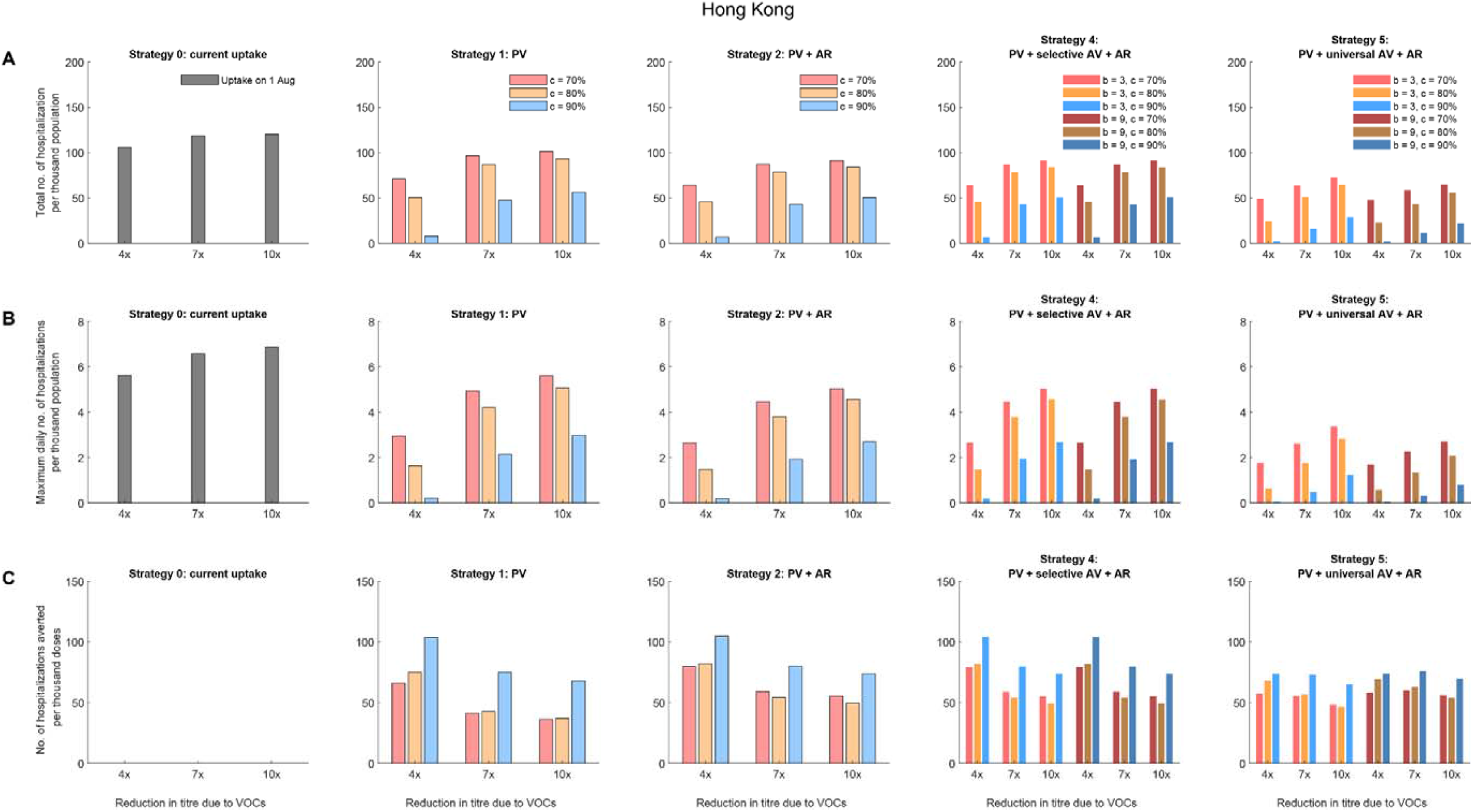
Comparative effectiveness of allocation strategies of vaccines and antivirals in Hong Kong assuming all vaccinees received the BNT162b2 vaccines. (A) The total number of hospitalizations per thousand population. (B) The maximum daily number of hospitalizations per thousand population. (C) The number of hospitalizations averted per thousand vaccine doses, compared with the existing vaccine uptake as of 1 August 2021 (i.e., 44.5%).

**Figure S5.**
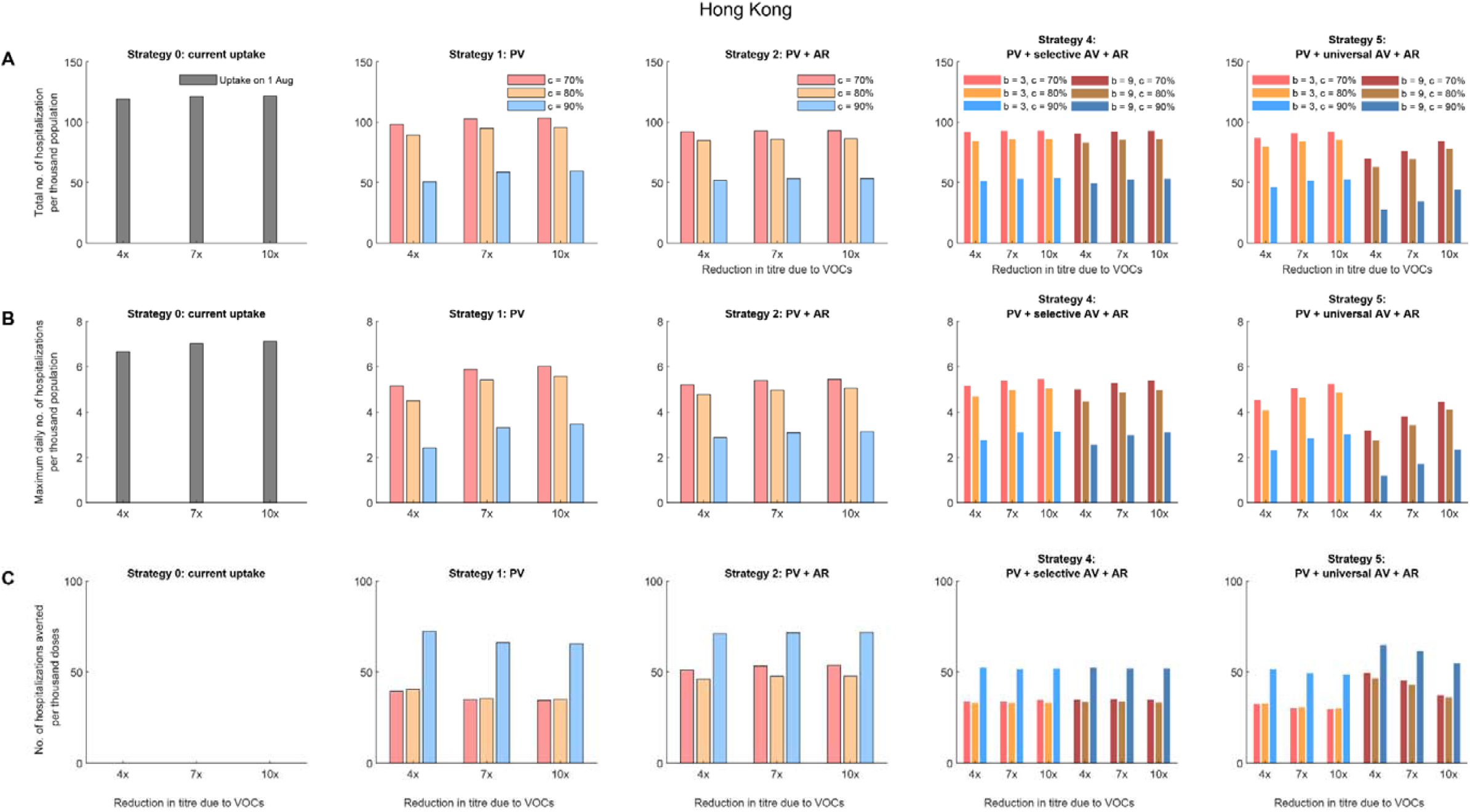
Comparative effectiveness of allocation strategies of vaccines and antivirals in Hong Kong assuming all vaccinees received the CoronaVac vaccines. (A) The total number of hospitalizations per thousand population. (B) The maximum daily number of hospitalizations per thousand population. (C) The number of hospitalizations averted per thousand vaccine doses, compared with the existing vaccine uptake as of 1 August 2021 (i.e., 44.5%).

**Figure S6.**
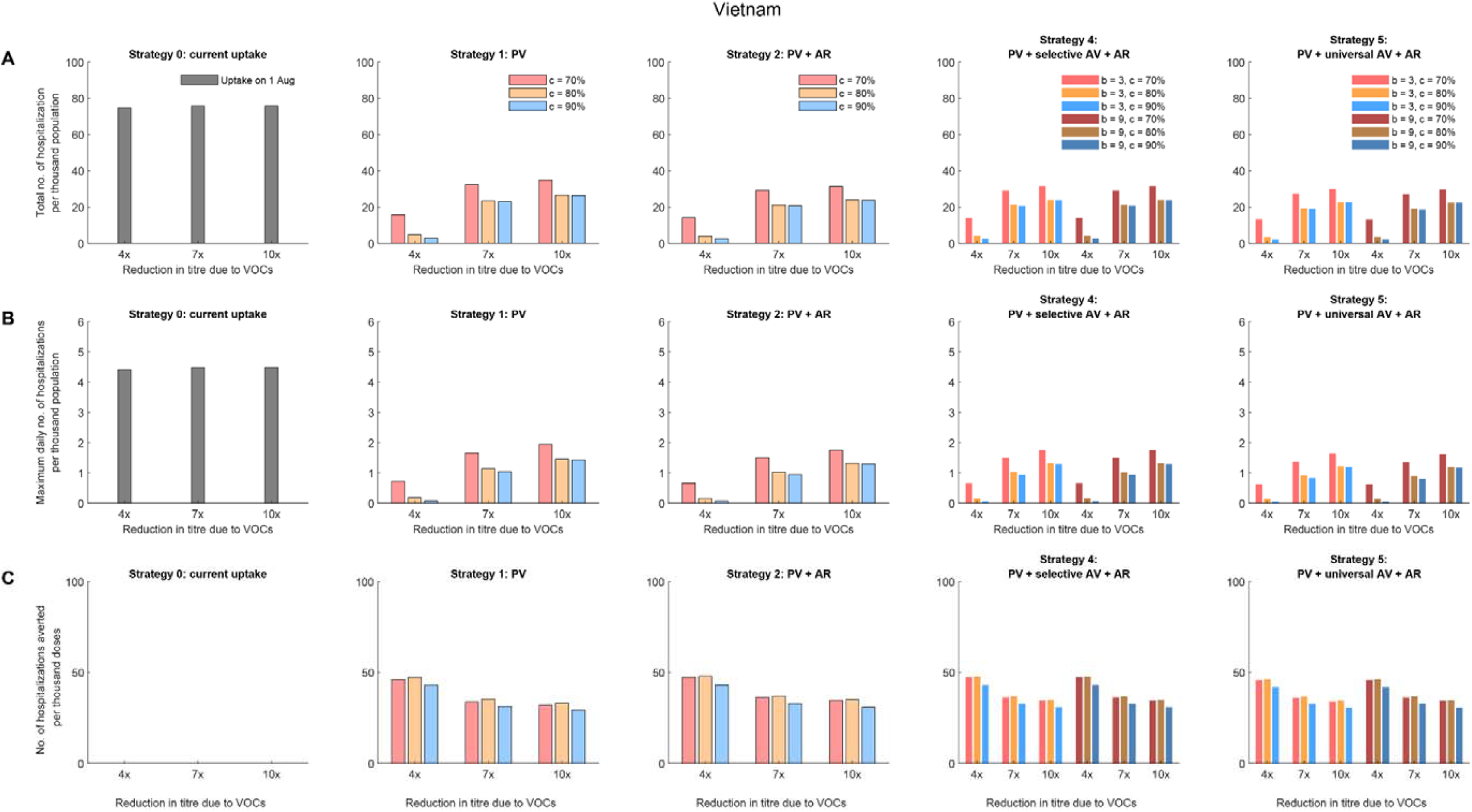
Comparative effectiveness of allocation strategies of vaccines and antivirals in Vietnam assuming all vaccinees received BNT162b2 vaccines. (A) The total number of hospitalizations per thousand population. (B) The maximum daily number of hospitalizations per thousand population. (C) The number of hospitalizations averted per thousand vaccine doses, compared with the existing vaccine uptake as of 1 August 2021 (i.e., about 6% coverage).

**Figure S7.**
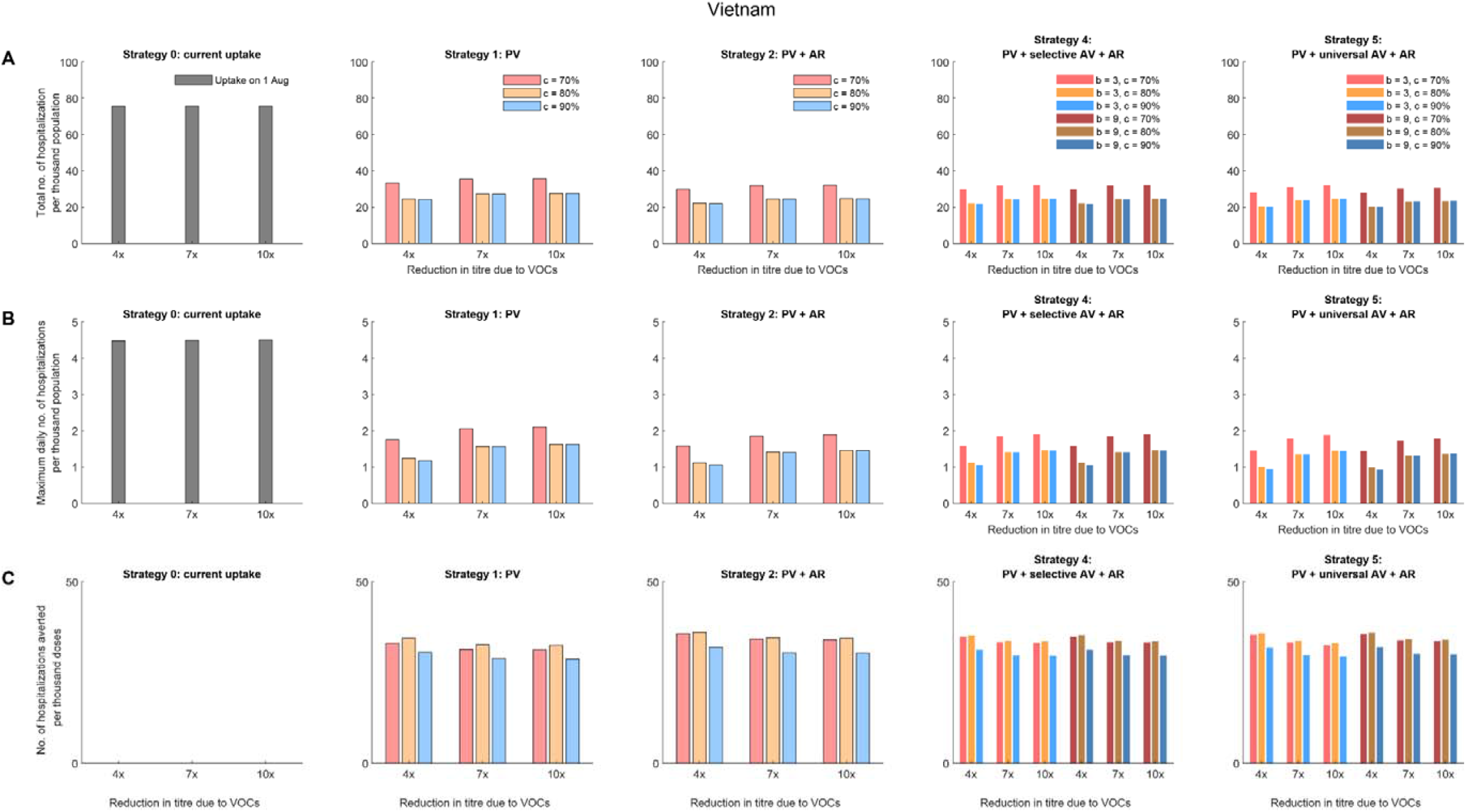
Comparative effectiveness of allocation strategies of vaccines and antivirals in Vietnam assuming all vaccinees received AZD1222 vaccines. (A) The total number of hospitalizations per thousand population. (B) The maximum daily number of hospitalizations per thousand population. (C) The number of hospitalizations averted per thousand vaccine doses, compared with the existing vaccine uptake as of 1 August 2021 (i.e., about 6% coverage).

**Figure S8.**
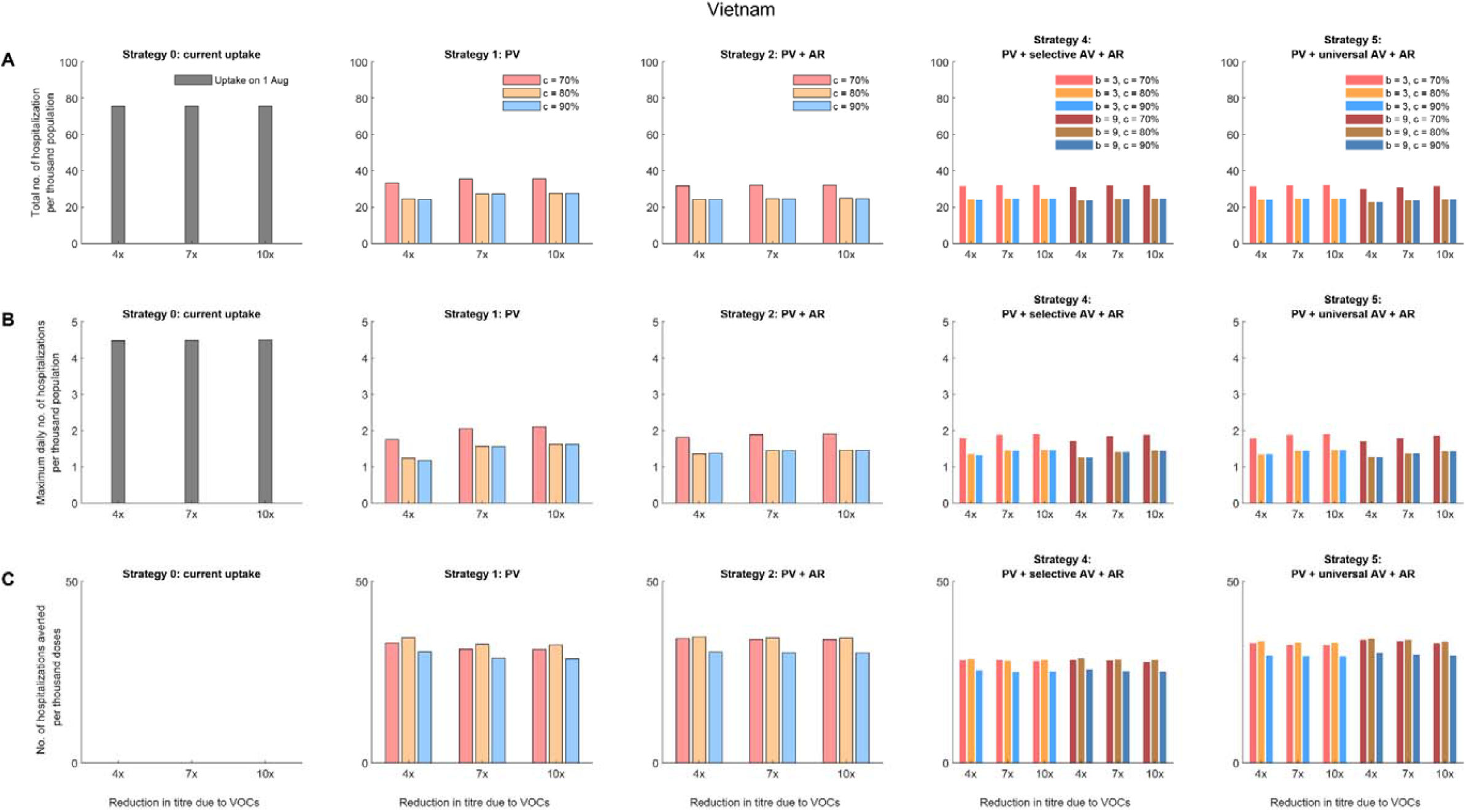
Comparative effectiveness of allocation strategies of vaccines and antivirals in Vietnam assuming all vaccinees received CoronaVac vaccines. (A) The total number of hospitalizations per thousand population. (B) The maximum daily number of hospitalizations per thousand population. (C) The number of hospitalizations averted per thousand vaccine doses, compared with the existing vaccine uptake as of 1 August 2021 (i.e., about 6% coverage).

**Table S1.** Age-specific vaccine uptake in Japan, Hong Kong, and Vietnam as of 1 August 2021.

| Age group | Japan | Hong Kong | Vietnam |
| --- | --- | --- | --- |
| 0-11 | 0% | 0% | 0% |
| 12-19 | 23.3% | 39% | 0% |
| 20-29 | 23.3% | 49% | 9.5% |
| 30-39 | 23.3% | 56% | 9.5% |
| 40-49 | 23.3% | 66% | 9.5% |
| 50-59 | 23.3% | 59% | 9.5% |
| 60-69 | 55.5% | 43% | 9.5% |
| 70-79 | 87.6% | 28% | 0% |
| 80 and above | 87.6% | 8% | 0% |

**Table S2.** Assumption about PRNT50 titre threshold and proportion of vaccinees with titres at or above the original threshold.

| PRNT50 titre threshold against the original virus ($r$) | Protection against infection of the original virus ($\epsilon_m(r)$) | Proportion of BNT162b2 vaccinees ≥ the original threshold ($F_b(r)$) | Proportion of CoronaVac vaccinees ≥ the original threshold ($F_c(r)$) |
| --- | --- | --- | --- |
| 25.9 | 0.5 | 100% (99-100) | 86.2% (85.7-86.7) |
| 39.9 | 0.6 | 100% (99-100) | 73.0% (72.3-73.6) |
| 54.4 | 0.7 | 99.7% (99.6-99.8) | 60.6% (60.0-61.3) |
| 74.1 | 0.8 | 98.7% (98.5-98.9) | 47.2% (46.5-47.9) |
| 99.7 | 0.9 | 95.5% (95.2-95.8) | 34.5% (33.8-35.2) |

**Table S3.**
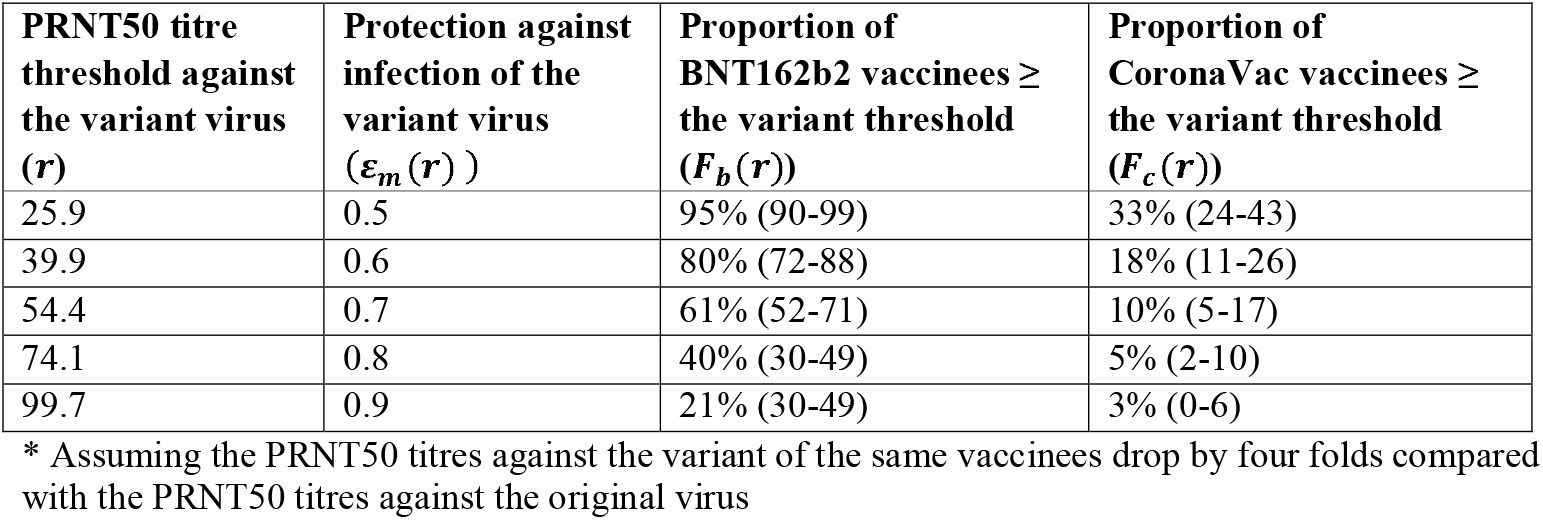
Assumption about PRNT50 titre threshold and proportion of vaccinees with titres at or above the variant threshold*.

**Table S4.** Model parameters.

| Parameter | Description, assumption, and source | Value |
| --- | --- | --- |
| $R_e$ | Effective reproductive number in the absence of vaccination, considering the emergence of variants of concern (assumed) | 6.0 |
| $T_{GT}$ | Mean generation time <sup>32</sup> | 5.4 days |
| $f_{GT}$ | Probability density function of generation time <sup>32</sup> | Gamma (4, 1.35) |
| $\sigma_m$ | Vaccine efficacy in reducing susceptibility | Estimated |
| $\sigma_t$ | Vaccine efficacy in reducing infectivity | Assumed to be $0.8 \times \sigma_m$ |
| $\sigma_s$ | Vaccine efficacy in reducing symptomatic diseases and hospitalizations | Assumed to be $1.25 \times \sigma_m$ and between 0.65 and 0.95 |
| $\varepsilon_m$ | Antiviral efficacy in reducing susceptibility | 0 |
| $\varepsilon_t$ | Antiviral efficacy in reducing infectivity | 0 |
| $\varepsilon_s$ | Antiviral efficacy in reducing symptomatic diseases and hospitalizations <sup>2</sup> | 0.5 |
| $p_{n,symptom}$ | The probability of developing symptomatic diseases if infected, for unvaccinated individuals (estimated from preliminary data from the Guangzhou Delta outbreak) | 60% |
| $p_{v,symptom}$ | The probability of developing symptomatic diseases if infected, for vaccinated individuals (estimated from preliminary data from the Guangzhou Delta outbreak) | 40% |
| $p_{a,death}$ | Age-specific infection fatality risk of a VOC similar to the Delta variant <sup>33,34</sup>; assuming the hazard ratio of Delta variant was 1.45 times of that of Alpha variant | Age 0-34: 0.044%<br>Age 35-54: 0.11%<br>Age 55-69: 0.86%<br>Age 70-84: 8.7%<br>Age $\geq 85$: 33% |
| $p_{a,hospitalization}$ | Age-specific infection hospitalization risk of a VOC similar to the Delta variant <sup>33,34</sup>; assuming the hazard ratio of Delta variant was 2.26 times of that of Alpha variant | Age 0-9: 0.0036%<br>Age 10-19: 0.0902%<br>Age 20-29: 2.35%<br>Age 30-39: 7.75%<br>Age 40-49: 9.83%<br>Age 50-59: 18.44%<br>Age 60-69: 26.67%<br>Age 70-79: 37.52%<br>Age $\geq 80$: 41.58% |
| $f_{incubation}$ | Probability density function of incubation period <sup>35</sup> | Lognormal distribution<br>Mean: 5.22 days<br>SD: 3.9 days |
| $f_{hospitalization}$ | Probability density function of the time between infection and hospitalization <sup>36</sup> | Gamma distribution<br>Mean: 8 days<br>SD: 3.6 days |
| $f_{death}$ | Probability density function of the time between infection and death; estimated from $f_{incubation}$ and the probability density function of the time between onset and death (Mean 18.8 days and SD 8.46 days) from Verity et al <sup>36</sup>; | Gamma distribution<br>Mean: 23.0 days<br>SD: 9.9 days |
| $H_{max}$ | The maximum number of COVID-19 hospitalizations that the local health system could take care of per day (assuming it is similar to the daily number of COVID-19 hospitalizations admitted in the UK in early Jan 2021 <sup>37</sup>; On 1 Jan 2021, the number of hospital admissions in the UK was 3,371; The maximum daily number of hospital admissions in the UK since the emergence of COVID-19 was 4,574 on 12 Jan 2021) | 0.005% of the total population |

## References

1. Ritchie H, Ortiz-Ospina E, Beltekian D, et al. Coronavirus pandemic (COVID-19). Our World in Data; 2021.

2. Merck. Merck and Ridgeback’s Investigational Oral Antiviral Molnupiravir Reduced the Risk of Hospitalization or Death by Approximately 50 Percent Compared to Placebo for Patients with Mild or Moderate COVID-19 in Positive Interim Analysis of Phase 3 Study. 2021. https://www.merck.com/news/merck-and-ridgebacks-investigational-oral-antiviral-molnupiravir-reduced-the-risk-of-hospitalization-or-death-by-approximately-50-percent-compared-to-placebo-for-patients-with-mild-or-moderat/.

3. Wadman M. Israel’s grim warning: Delta can overwhelm shots. American Association for the Advancement of Science; 2021.

4. Lucas C, Vogels CBF, Yildirim I, et al. Impact of circulating SARS-CoV-2 variants on mRNA vaccine-induced immunity in uninfected and previously infected individuals. Nature 2021.

5. Lopez Bernal J, Andrews N, Gower C, et al. Effectiveness of Covid-19 Vaccines against the B.1.617.2 (Delta) Variant. New England Journal of Medicine 2021; 385(7): 585–94.

6. Puranik A, Lenehan PJ, Silvert E, et al. Comparison of two highly-effective mRNA vaccines for COVID-19 during periods of Alpha and Delta variant prevalence. medRxiv 2021: 2021.08.06.21261707.

7. Leung K, Wu JT, Leung GM. Effects of adjusting public health, travel, and social measures during the roll-out of COVID-19 vaccination: a modelling study. The Lancet Public Health 2021.

8. Tregoning JS, Flight KE, Higham SL, Wang Z, Pierce BF. Progress of the COVID-19 vaccine effort: viruses, vaccines and variants versus efficacy, effectiveness and escape. Nature Reviews Immunology 2021.

9. Mok CKP, Cohen CA, Cheng S, et al. Comparison of the Immunogenicity of BNT162b2 and CoronaVac COVID-19 Vaccines in Hong Kong: An Observational Cohort Study. Available at SSRN: https://ssrncom/abstract=3884943 or http://dxdoiorg/102139/ssrn3884943 2021.

10. Feng S, Phillips DJ, White T, et al. Correlates of protection against symptomatic and asymptomatic SARS-CoV-2 infection. Nature Medicine 2021.

11. Krause PR, Fleming TR, Peto R, et al. Considerations in boosting COVID-19 vaccine immune responses. The Lancet.

12. Tang P, Hasan MR, Chemaitelly H, et al. BNT162b2 and mRNA-1273 COVID-19 vaccine effectiveness against the Delta (B.1.617.2) variant in Qatar. medRxiv 2021: 2021.08.11.21261885.

13. Tartof SY, Slezak JM, Fischer H, et al. Effectiveness of mRNA BNT162b2 COVID-19 vaccine up to 6 months in a large integrated health system in the USA: a retrospective cohort study. The Lancet.

14. Goel RR, Painter MM, Apostolidis SA, et al. mRNA vaccines induce durable immune memory to SARS-CoV-2 and variants of concern. Science; 0(0): eabm0829.

15. Laurie MT, Liu J, Sunshine S, et al. Exposures to different SARS-CoV-2 spike variants elicit neutralizing antibody responses with differential specificity towards established and emerging strains. medRxiv 2021: 2021.09.08.21263095.

16. Cevik M, Grubaugh ND, Iwasaki A, Openshaw P. COVID-19 vaccines: Keeping pace with SARS-CoV-2 variants. Cell.

17. Khoury DS, Cromer D, Reynaldi A, et al. Neutralizing antibody levels are highly predictive of immune protection from symptomatic SARS-CoV-2 infection. Nature medicine 2021: 1-7.

18. Sadarangani M, Marchant A, Kollmann TR. Immunological mechanisms of vaccine-induced protection against COVID-19 in humans. Nature Reviews Immunology 2021: 1–10.

19. Geers D, Shamier MC, Bogers S, et al. SARS-CoV-2 variants of concern partially escape humoral but not T cell responses in COVID-19 convalescent donors and vaccine recipients. Science Immunology 2021; 6(59): eabj1750.

20. Mateus J, Dan JM, Zhang Z, et al. Low-dose mRNA-1273 COVID-19 vaccine generates durable memory enhanced by cross-reactive T cells. Science; 0(0): eabj9853.

21. Scientific Advisory Group for Emergencies. SPI-M-O: Summary of further modelling of easing restrictions – Roadmap Step 4 on 19 July 2021, 7 July 2021. 2021. https://assets.publishing.service.gov.uk/government/uploads/system/uploads/attachment_data/file/1001169/S1301_SPI-M-O_Summary_Roadmap_second_Step_4.21_.pdf.

22. Wu JT, Leung K, Bushman M, et al. Estimating clinical severity of COVID-19 from the transmission dynamics in Wuhan, China. Nature medicine 2020; 26(4): 506–10.

23. Falsey AR, Frenck RW, Walsh EE, et al. SARS-CoV-2 Neutralization with BNT162b2 Vaccine Dose 3. New England Journal of Medicine 2021.

24. Flaxman A, Marchevsky NG, Jenkin D, et al. Reactogenicity and immunogenicity after a late second dose or a third dose of ChAdOx1 nCoV-19 in the UK: a substudy of two randomised controlled trials (COV001 and COV002). The Lancet 2021; 398(10304): 981–90.

25. Pan H, Wu Q, Zeng G, et al. Immunogenicity and safety of a third dose, and immune persistence of CoronaVac vaccine in healthy adults aged 18-59 years: interim results from a double-blind, randomized, placebo-controlled phase 2 clinical trial. medRxiv 2021: 2021.07.23.21261026.

26. Li M, Yang J, Wang L, et al. A booster dose is immunogenic and will be needed for older adults who have completed two doses vaccination with CoronaVac: a randomised, double-blind, placebo-controlled, phase 1/2 clinical trial. medRxiv 2021: 2021.08.03.21261544.

27. Bonelli M, Mrak D, Tobudic S, et al. Additional heterologous versus homologous booster vaccination in immunosuppressed patients without SARS-CoV-2 antibody seroconversion after primary mRNA vaccination: a randomized controlled trial. medRxiv 2021: 2021.09.05.21263125.

28. Dhar MS, Marwal R, Radhakrishnan V, et al. Genomic characterization and Epidemiology of an emerging SARS-CoV-2 variant in Delhi, India. medRxiv 2021: 2021.06.02.21258076.

29. Reynolds σ, Pade C, Gibbons JM, et al. Prior SARS-CoV-2 infection rescues B and T cell responses to variants after first vaccine dose. Science 2021; 372(6549): 1418–23.

30. The Government of Hong Kong Special Administrative Region. Hong Kong Vaccination Dashboard. 2021. https://www.covidvaccine.gov.hk/en/dashboard.

31. Lau EH, Tsang OT, Hui DS, et al. Neutralizing antibody titres in SARS-CoV-2 infections. Nature communications 2021; 12(1): 1–7.

32. Leung K, Wu JT, Liu D, Leung GM. First-wave COVID-19 transmissibility and severity in China outside Hubei after control measures, and second-wave scenario planning: a modelling impact assessment. The Lancet 2020.

33. Davies NG, Jarvis CI, Edmunds WJ, Jewell NP, Diaz-Ordaz K, Keogh RH. Increased mortality in community-tested cases of SARS-CoV-2 lineage B. 1.1. 7. Nature 2021; 593(7858): 270–4.

34. Twohig KA, Nyberg T, Zaidi A, et al. Hospital admission and emergency care attendance risk for SARS-CoV-2 delta (B.1.617.2) compared with alpha (B.1.1.7) variants of concern: a cohort study. The Lancet Infectious Diseases.

35. Li Q, Guan X, Wu P, et al. Early Transmission Dynamics in Wuhan, China, of Novel Coronavirus–Infected Pneumonia. New England Journal of Medicine 2020.

36. Verity R, Okell LC, Dorigatti I, et al. Estimates of the severity of coronavirus disease 2019: a model-based analysis. The Lancet Infectious Diseases 2020; 20(6): 669–77.

37. GOV.UK. Coronavirus (COVID-19) in the UK: Healthcare in United Kingdom. 2021. https://coronavirus.data.gov.uk/details/healthcare.

